# Accuracy of clinical risk factor-based models as a screening test for detecting gestational diabetes mellitus in a low-resource setting

**DOI:** 10.1101/2023.07.14.23292690

**Authors:** Olayinka Comfort Senbanjo, Fatimat Motunrayo Akinlusi, Kabiru Afolarin Rabiu

## Abstract

Current World Health Organization guidelines recommend fasting 2-hour tests for all pregnant women, a strategy that is burdensome for patients as well as time and labor-intensive for health systems. There have been suggestions for the use of clinical risk factors-based models as alternatives. These have not been widely tested especially in low-resource countries. We aimed to determine the prevalence of GDM and the accuracy of clinical risk factor-based models as screening tools for detecting GDM.

This was a prospective cohort study of consenting 400 pregnant women receiving antenatal care at a tertiary health facility in Lagos, Nigeria. All the study subjects were assessed for the risk of GDM using three different clinical risk-based models. They also had universal screening for GDM at 24–28 weeks gestational age using the gold standard 2-hour 75g Oral Glucose Tolerance Test (OGTT). Statistical analysis was done using the statistical package for social science version 24. The Receiver Operating Curve (ROC) was used to determine the accuracy of the risk factor-based models.

The mean age of the subjects was 31.0 **±** 5.3 years. A total of 76 subjects met the IADPSG/WHO 2013 criteria giving a prevalence of 19.0%. According to the clinical risk scores by Naylor *et al*, Caliskan *et al* and Phaloprakarn *et al,* 340 (85%), 269 (67.3%) and 375 (93.8%) participants respectively had a risk score positive for GDM. If the study participants were selectively screened based on these models, between 71.1-96.1% of the women with GDM would have been identified and 6.3-32.8% of the women would not have performed the diagnostic test. The models had areas under the ROC that ranges between 51.6-52.9%.

**Conclusions:** The prevalence of GDM is high and the clinical risk factor-based prediction models tested in this study could be used to stratify low-risk women out of diagnostic tests.

## Introduction

Globally, the prevalence of gestational diabetes mellitus varies between 1-28% depending on the location, characteristics of the studied population, and the diagnostic criteria used.^1^ More than 80% of the burden of GDM is found in low- and middle-income countries and it is speculated to contribute significantly to the high maternal and infant mortality rates in these countries.^2^ In the United States^3^, recent estimates show that up to 9% of all pregnancies are complicated by GDM while the mean prevalence of GDM for countries in Europe is 5.4%.^4^ In Africa, a systematic review of studies on GDM reported pooled prevalence of about 14%, contributing significantly to the total global burden of gestational diabetes.^5^ In Nigeria, the prevalence of GDM ranges between 0.3% and 35.9%.^6, 7^ It is reported that Nigeria has the highest prevalence in Africa and GDM is one of the five commonest medical conditions complicating pregnancy in women attending maternal and child health care facilities in Nigeria. ^7–9^

It is important that every pregnant woman must be screened for GDM because pregnancy presents a window period during which it can be identified and treated to ensure a good pregnancy outcome and as well halt the progression to Type 2 diabetes mellitus and other severe consequences.

To date, no consensus has been reached worldwide regarding the best screening method for the identification of GDM nor the best diagnostic criteria for an accurate diagnosis for GDM.^10^ The methods of screening GDM and the diagnostic criteria used vary from country to country and from centre to centre.^10^ The various screening methods used for GDM include assessment of random blood sugar, fasting blood sugar, glucose in the urine, clinical risk factors, and Oral Glucose Challenge Test (GCT) while the diagnosis of GDM is confirmed by the Oral Glucose Tolerance Test (OGTT) which is considered the gold standard.^10^

The American College of Obstetrics and Gynaecology recommends a universal screening approach that involves screening all pregnant women for GDM by the use of a 50g glucose challenge test.^11^ The universal method of screening has been adopted by obstetricians both in developed and developing countries.^12^ Although this method will enable the health care providers to pick up almost all the affected pregnant women with GDM, this approach may be impracticable in low-and middle-income countries due to cost implications and therefore low acceptance of the test.^12^

GDM is known to be associated with some risk factors. As a result, the fourth International Workshop-Conference on GDM recommended a selective screening method whereby those women with certain clinical characteristics are selected and thereafter offered Oral Glucose Tolerance Test (OGTT) to diagnose GDM.^13^ In sub-Sahara African countries, many health facilities relied on a checklist of these identified risk factors to select pregnant women who should undergo the diagnostic test using the Oral Glucose Tolerance Test (OGTT).^8, 14, 15^ However, GDM in this high-risk population remained under-diagnosed.^8, 15^ The use of risk factors as a screening test and the diagnostic accuracy of the risk factors is unclear either when used separately or collectively as a checklist. It is also unknown whether the use of the risk factor assessment is preferable to plasma glucose measurement as a general screening strategy for GDM at the first contact. There are established clinical risk factor-based models that have been developed and advocated for use in some countries such as Naylor et al in Canada,^16^ Caliskan in Turkey^17^, and Phaloprakarn in Bangladesh.^18^ The widespread applicability of these models has not been tested especially in low-and middle-income countries.

The aim of this study was to determine the incidence of GDM and to evaluate the diagnostic accuracy of these established clinical risk factor-based models for the detection of GDM in Lagos, Nigeria.

## Materials and methods

### Study setting

The study was carried out at the antenatal clinic of the Institute of Maternal and Child Health, Ayinke House, Lagos State University Teaching Hospital (LASUTH), Ikeja, Lagos State, South-Western, Nigeria between 8th March 2021 and 6^th^ December 2021. LASUTH is one of the two teaching hospitals in Lagos State. The hospital is located in Ikeja Local Government Area and it is owned by the Lagos State Government. It serves the purpose of a training center for resident Doctors as well as providing health care for the people of Lagos state and its environment. The Institute of Maternal and Child Health, Ayinke House, is run by the Department of Obstetrics and Gynaecology. The obstetric arm is run via the antenatal clinic as well as via the emergency room. The obstetric arm provides care for both booked and un-booked pregnant women. The ANC receives and cares for all pregnant women that decide to book in LASUTH. ANC runs every day of the week excluding weekends with an average of 30 patients daily. The ANC is run by an average of four Consultant Obstetricians and Gynaecologists, five Resident doctors, and four nurses.

### Study design

This study was a prospective, hospital-based, cohort study of all consecutive consenting pregnant women at the gestational age of 24 weeks to 28 weeks.

### Inclusion criteria

These are pregnant women that booked for antenatal care and had their booking weight and height measured and recorded at gestational age less than or equal to 24 weeks and with singleton pregnancy.

### Exclusion criteria

Pregnant women with a history of pre-gestational diabetes mellitus, on drugs that can affect glycaemic profile such as steroids and beta-agonist, multiple pregnancies, and unwilling to participate in the study were excluded from the study.

### Ethical approval

Approval to conduct this study was obtained from the Health Research and Ethics Committee of the Lagos State University Teaching Hospital, Ikeja, Lagos State with approval number LREC/06/10/1403. Written informed consent was obtained from participants who agree to participate in the study.

### Sample size

The minimum number of subjects ‘n’ required for the study was estimated from the formula:

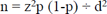

Where ‘n’ is the desired sample size,

‘z’ is the critical value and in a two-tailed test, it is equal to 1.96.

‘p’ is the prevalence of gestational diabetes from previous studies in Nigeria. Previous studies in Nigeria gave the prevalence of gestational diabetes within ranges of 0.5% - 35.9%. For this study, a prevalence of 35.9% by Onyenekwe *et al* was used.^7^

‘d’ is the absolute sampling error that can be tolerated. In this study, it was fixed at 5 percent

Therefore, the minimum sample size ‘n’ = 1.96^2^ x 0.359 x (1 – 0.359) ÷ 0.05^2^ = <u>353.8</u> which is approximately 354. Taking into consideration a possible attrition rate of 10% among pregnant women, the minimum sample size for this study was <u>389.4.</u> This sample size was rounded up to 400.

### Selection of study participants and test procedures

Prior to the commencement of the study, the researcher informed the doctors and nurses in the Department of Obstetrics and Gynaecology about the research and the recruitment protocol. The researcher also trained three research assistants on the use of the study proforma and the different measurements to be taken with hands-on demonstration to ensure standardization.

Data were obtained from all consented pregnant women using a purposely designed interviewer-administered questionnaire and the information in Naylor et al,^16^ Caliskan et al,^17^ and Phalopraskan et al risk scoring models.^18^ Information obtained includes socio-demographic characteristics, obstetric history (such as parity, previous miscarriage, history of GDM, history of perinatal death), and family history of diabetes mellitus. The families were assigned a socio-economic class using the method recommended by Ogunlesi *et al*^19^. Those with mean scores of 1 and 2 were further classified as upper class, those with mean scores of 3 were classified as middle class, while those with mean scores of 4 and 5 were classified as falling in the lower social class. Dating of the pregnancy and gestational age estimation was done by the use of the first day of the last menstrual period in a woman with a 28-day menstrual cycle and this was compared with the pregnant woman’s fundal height. The researcher used the earliest ultrasound (i.e. a scan done at less or equal to the estimated gestational age of 20 weeks) if the subject was not sure of her date. The weight and height of all subjects measured at gestational age less than or equal to 24 weeks were recorded. Random blood glucose measurements were done in all selected subjects at the first contact. Thereafter, all the selected subjects were requested to return to the antenatal clinic fasting after a week for Oral Glucose Tolerance Test (OGTT) to confirm the presence of GDM. The test was performed between the 24^th^ and 28^th^ week of gestation.

A venous blood sample was obtained from the participants to perform random blood glucose tests using an analyzer that operates on the principle of the enzymatic oxidase-peroxidase method of glucose estimation. After ensuring aseptic procedure, about three milliliters of venous blood was drawn from a vein in the antecubital fossa using a 21G needle butterfly device with a safety system. The blood was collected into a vacutainer tube containing the glycolytic inhibitor sodium fluoride and kept on ice from the time of phlebotomy until delivery to the laboratory. Each blood sample was centrifuged to obtain plasma within two to four hours of conducting phlebotomy. The plasma samples were stored at a temperature of −20^0^c and pooled together to determine plasma glucose concentration. The value obtained was recorded in the participant’s study proforma.

All study participants were to present in the early morning for an oral glucose tolerance test following a period of about 8-12 hour overnight fast. Each patient was telephoned the day before the test to remind her to fast. A 75-gram glucose load was prepared by dissolving 75g of anhydrous glucose in 250-300 ml of clean water. The participants were advised to consume the liquid as quickly as is comfortable for them and to remain relaxed and avoid vigorous activity during the test. Venous blood for glucose measurement was collected as described above and samples were analyzed using the enzymatic reaction of oxidase-peroxidase method of glucose estimation.

The first sample collected prior to the glucose load is the fasting plasma glucose, while the second and third blood samples shall be taken at 1 hour and 2 hours respectively after the glucose load.

### Diagnostic cut-offs

Gestational diabetes mellitus in the participants was diagnosed using the criteria of the clinical risk factor-based models and the plasma glucose values according to the International Association of Diabetes and Pregnancy Study Groups/World Health Organization (IADPSG/WHO 2013) criteria.^10, 12^ GDM prevalence as determined by IADPSG/WHO 2013 criteria was used as the standard reference.

#### Naylor et al clinical risk score^16^

The screening score is based on the clinical variables of age, Body Mass Index (BMI) and ethnicity. Based on these variables, women were assigned a clinical risk score, with a maximum obtainable score of 10 points. Women with scores of 0-1 were categorized as low risk while those with scores of 2-3 and those with scores higher than 3 are categorized as intermediate and high risk respectively. For this study, those with a score of 0-1 were considered negative for GDM while those with a score of 2-10 were considered positive for GDM.

#### Caliskan et al clinical risk score^17^

The screening score was based on clinical variables of age, BMI, family history of diabetes mellitus, a previous baby with birth weight greater than 4000g, previous adverse pregnancy outcome (defined as the presence of any of the following; recurrent spontaneous abortions, fetal anomaly despite a normal karyotype and prior unexplained in utero fetal death at a gestational age ≥ 20 weeks). A score of one was assigned for the presence of each of the five variables making a maximum obtainable core of five. A risk score of ≥ 2 is considered sufficient to diagnose GDM.

#### Phaloprakarn et al clinical risk score^18^

This risk factor-based model was based on clinical risk variables of age greater than 35 years, BMI greater than 27kg/m^2^, any first-degree relatives with type 2 diabetes mellitus or personal history of GDM, prior delivery of macrosomic infant, previous adverse pregnancy outcome (≥ 2 miscarriages, congenital malformation or stillbirth). The risk score is derived from the equation: 6 multiplied by the age of the woman + 11 multiplied by the BMI + 109 (If there is a family history of diabetes in a first-degree relative) + 42 (If there is prior delivery of a baby with birth weight greater than 4000 g) + 49 (If there is adverse pregnancy outcome such as 2 or more abortions, congenital malformation and stillbirth. A score of ≥ 380 is a positive screen for GDM.

### Data analysis

Data entry and analysis were done using the Statistical Package for Social Sciences (SPSS) version 24 software. Descriptive statistics such as frequency, percentages, means, standard deviation and the corresponding 95% CI were used to summarize the variables. The sensitivity, specificity, positive predictive and negative predictive values of the clinical risk factor-based models were calculated by determining the total number of women with true positive, true negative, false positive and false negative results with reference to the gold standard 2-hour 75g OGTT. A receiver-operating characteristics (ROC) curve was plotted and the area under the curve (AUC) was calculated to determine the overall test performance of the clinical risk factor-based models. A p-value of less than 0.05 was considered statistically significant for all tests.

## Results

### Socio-demographic characteristics of the study participants

The socio-demographic characteristics of the participants are shown in Table I. Their age ranges from 18-51 years with a mean age of 31.0 **±** 5.3 years. Most (93.8%) of the participants were in the age group 21-40 years. They are predominantly of the Yoruba tribe (70%) and the modal parity was nullipara (53%). Three hundred and forty-five (86.3%) participants belonged to the high socio-economic class (social class 1).

**Table 1:** Socio-demographic and anthropometric characteristics of the study population.

| **Parameters** | **Frequency** | **Percentages** |
| --- | --- | --- |
| **Age (years)** |  |  |
| ≤ 20 | 5 | 1.2 |
| 21-30 | 196 | 49.0 |
| 31-40 | 179 | 44.8 |
| >40 | 20 | 5.0 |
| **Tribe** |  |  |
| Yoruba | 280 | 70.0 |
| Hausa | 1 | 0.2 |
| Ibo | 61 | 15.3 |
| Others | 58 | 14.5 |
| **Parity** |  |  |
| 0 | 212 | 53.0 |
| 1 | 98 | 24.5 |
| 2-4 | 81 | 20.3 |
| ≥5 | 9 | 2.2 |
| **Occupation** |  |  |
| Professional/technical/managerial | 145 | 36.2 |
| Clerical, sales and services, skilled manual | 232 | 58.0 |
| Unskilled manual, farmer, and other | 23 | 5.8 |
| **Highest level of education** |  |  |
| No formal education | 0 | 0.0 |
| Primary | 6 | 1.5 |
| Secondary | 57 | 14.2 |
| Tertiary | 337 | 84.3 |
| **Husband's Occupation** |  |  |
| Professional/technical/managerial | 258 | 64.5 |
| Clerical, sales and services, skilled manual | 136 | 34.0 |
| Unskilled manual, farmer, and other | 6 | 1.5 |
| **Husband's highest level of education** |  |  |
| No formal education | 1 | 0.2 |
| Primary | 4 | 1.0 |
| Secondary | 49 | 12.3 |
| Tertiary | 346 | 86.5 |
| **Social class** |  |  |
| 1 | 345 | 86.25 |
| 2 | 50 | 12.5 |
| 3 | 5 | 1.25 |
| **\*Weight (Kg)** | 72.9 ± 16.1 |  |
| **\*Height (Meter)** | 1.62 ± 0.065 |  |
| **Body mass index** |  |  |
| <18.5 | 10 | 2.5 |
| 18.5 -24.9 | 137 | 34.3 |
| 25-29.9 | 126 | 31.5 |
| ≥30 | 127 | 31.8 |

\*Values are means (SD)

### Obstetric characteristics of the study participants

Table II shows the obstetric history of the study participants. Thirty (7.5%) participants gave a history of previous delivery of a macrosomic baby, 22 (5.5%) had a previous history of stillbirth, 58 (14.5%) had a previous history of miscarriage, 23 (5.8%) had a previous history of GDM, 10 (2.5%) had a previous history of congenital anomaly and only one (0.25%) had a previous history of polyhydramnios. Fifteen (3.8%) of the participants had the conception of the index pregnancy by artificial reproductive technology.

**Table II:**
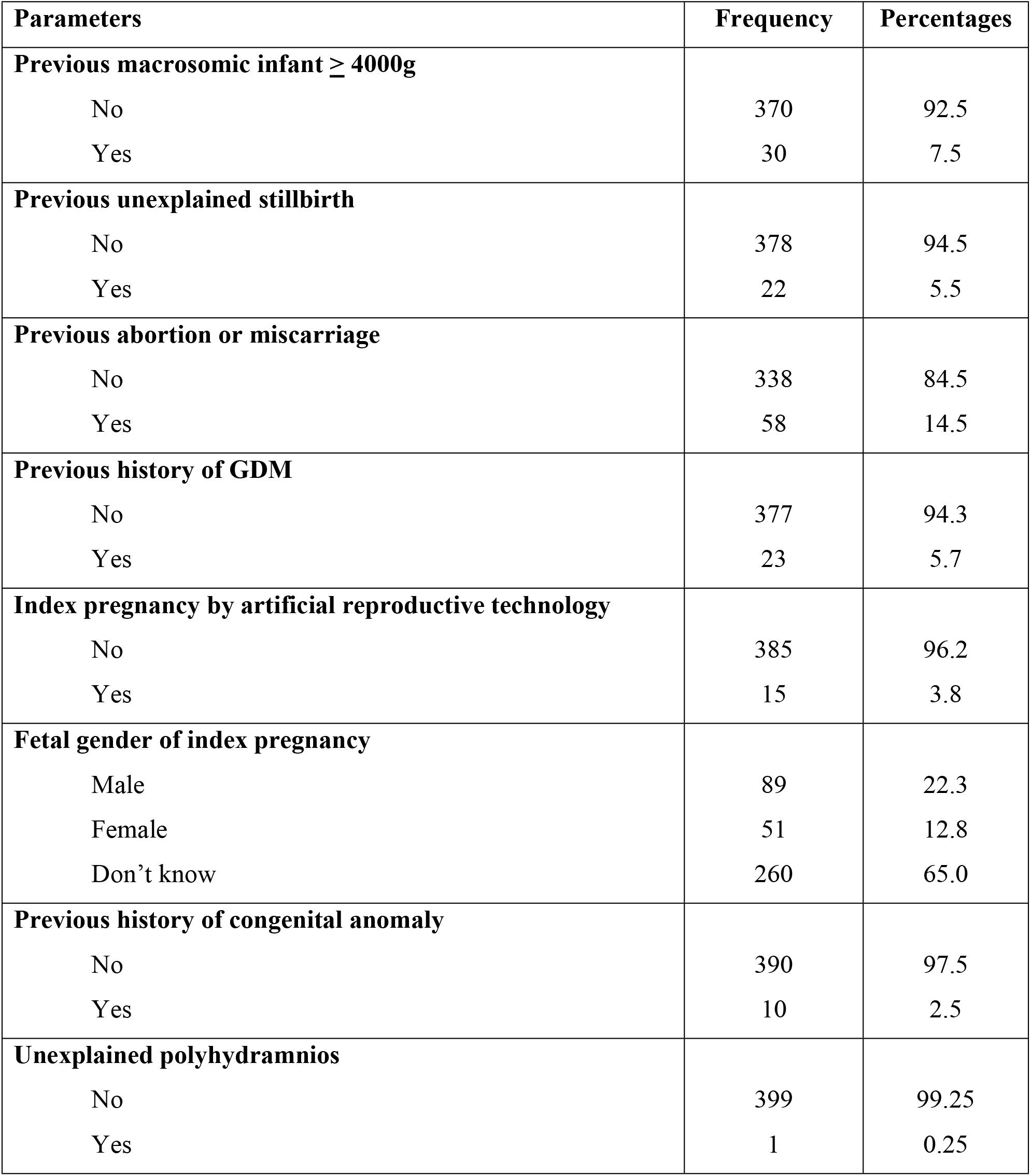
Obstetric characteristics of the study population.

| **Parameters** | **Frequency** | **Percentages** |
| --- | --- | --- |
| **Previous macrosomic infant $\geq 4000\text{g}$** |  |  |
| No | 370 | 92.5 |
| Yes | 30 | 7.5 |
| **Previous unexplained stillbirth** |  |  |
| No | 378 | 94.5 |
| Yes | 22 | 5.5 |
| **Previous abortion or miscarriage** |  |  |
| No | 338 | 84.5 |
| Yes | 58 | 14.5 |
| **Previous history of GDM** |  |  |
| No | 377 | 94.3 |
| Yes | 23 | 5.7 |
| **Index pregnancy by artificial reproductive technology** |  |  |
| No | 385 | 96.2 |
| Yes | 15 | 3.8 |
| **Fetal gender of index pregnancy** |  |  |
| Male | 89 | 22.3 |
| Female | 51 | 12.8 |
| Don't know | 260 | 65.0 |
| **Previous history of congenital anomaly** |  |  |
| No | 390 | 97.5 |
| Yes | 10 | 2.5 |
| **Unexplained polyhydramnios** |  |  |
| No | 399 | 99.25 |
| Yes | 1 | 0.25 |

### Prevalence of GDM using the various diagnostic criteria

Table III shows the prevalence of GDM based on various diagnostic and screening criteria. According to the IADPSG/WHO 2013 guideline, the prevalence of GDM based on the “gold standard” 2-hour OGTT was 13.0%. The prevalence based on FPG was 10.5%, whereas none of the participants met the criteria for GDM based on 1-hour OGTT. A total of 76 participants met the criteria of either 2-hour OGTT and or FPG giving a prevalence of 19.0%.

**Table III:**
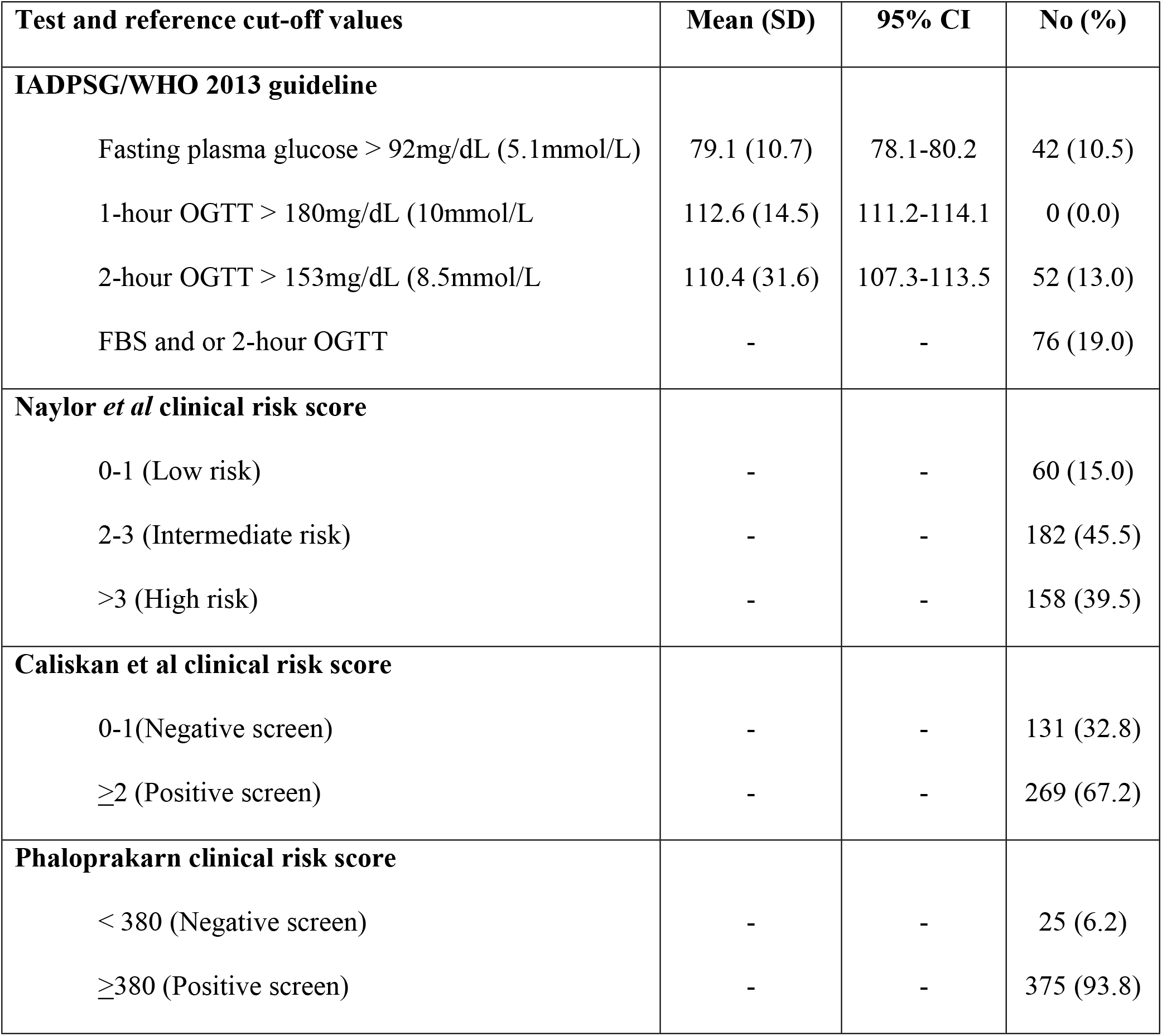
Prevalence of GDM using various diagnostic criteria.

| Test and reference cut-off values | Mean (SD) | 95% CI | No (%) |
| --- | --- | --- | --- |
| **IADPSG/WHO 2013 guideline** |  |  |  |
| Fasting plasma glucose > 92mg/dL (5.1mmol/L) | 79.1 (10.7) | 78.1-80.2 | 42 (10.5) |
| 1-hour OGTT > 180mg/dL (10mmol/L) | 112.6 (14.5) | 111.2-114.1 | 0 (0.0) |
| 2-hour OGTT > 153mg/dL (8.5mmol/L) | 110.4 (31.6) | 107.3-113.5 | 52 (13.0) |
| FBS and or 2-hour OGTT | - | - | 76 (19.0) |
| **Naylor *et al* clinical risk score** |  |  |  |
| 0-1 (Low risk) | - | - | 60 (15.0) |
| 2-3 (Intermediate risk) | - | - | 182 (45.5) |
| >3 (High risk) | - | - | 158 (39.5) |
| **Caliskan *et al* clinical risk score** |  |  |  |
| 0-1(Negative screen) | - | - | 131 (32.8) |
| ≥2 (Positive screen) | - | - | 269 (67.2) |
| **Phaloprakarn clinical risk score** |  |  |  |
| < 380 (Negative screen) | - | - | 25 (6.2) |
| ≥380 (Positive screen) | - | - | 375 (93.8) |

According to the clinical risk score by Naylor *et al*, 340 (85%) participants had a risk score that was significant enough to consider diagnostic screening for GDM while the risk score by Caliskan *et al* shows that 269 (67.3%) participants can be considered to have GDM. However, the clinical risk score by Phaloprakarn *et al* shows that 375 participants can be considered to have GDM.

### Accuracy of selected published clinical risk-based score for the diagnosis of GDM

Table IV shows the diagnostic accuracy of the selected published clinical risk score in the diagnosis of GDM using IADPSG/WHO 2013 diagnostic criteria as the gold standard. All the clinical risk scores tested have low specificity (6.7-33.6%) and PPV (19.5-20-1%). The accuracy was highest with the Caliskan clinical risk score screening test (40.8%).

**Table IV:** Accuracy of selected published clinical risk score in the diagnosis of GDM using IADPG/WHO 2013 diagnostic criteria as reference standard.

| **Parameter** | **TP** | **FN** | **FP** | **TN** | **Sensitivity**<br>**(%)** | **Specificity**<br>**(%)** | **PPV**<br>**(%)** | **NPV**<br>**(%)** | **Accuracy**<br>**(%)** |
| --- | --- | --- | --- | --- | --- | --- | --- | --- | --- |
| **Naylor *et al*** | 67 | 9 | 273 | 51 | 88.2 | 15.7 | 19.7 | 85.0 | 29.5 |
| **Caliskan *et al*** | 54 | 22 | 215 | 109 | 71.1 | 33.6 | 20.1 | 83.2 | 40.8 |
| **Phaloprakarn *et al*** | 73 | 3 | 302 | 22 | 96.1 | 6.7 | 19.5 | 88.0 | 23.8 |

Figures 1-3 shows the ROC curves of the clinical risk factor-based models for our population had AUC ranging between 51.6% to 52.9% indicating unsatisfactory discriminatory capacity.

**Figure 1:**
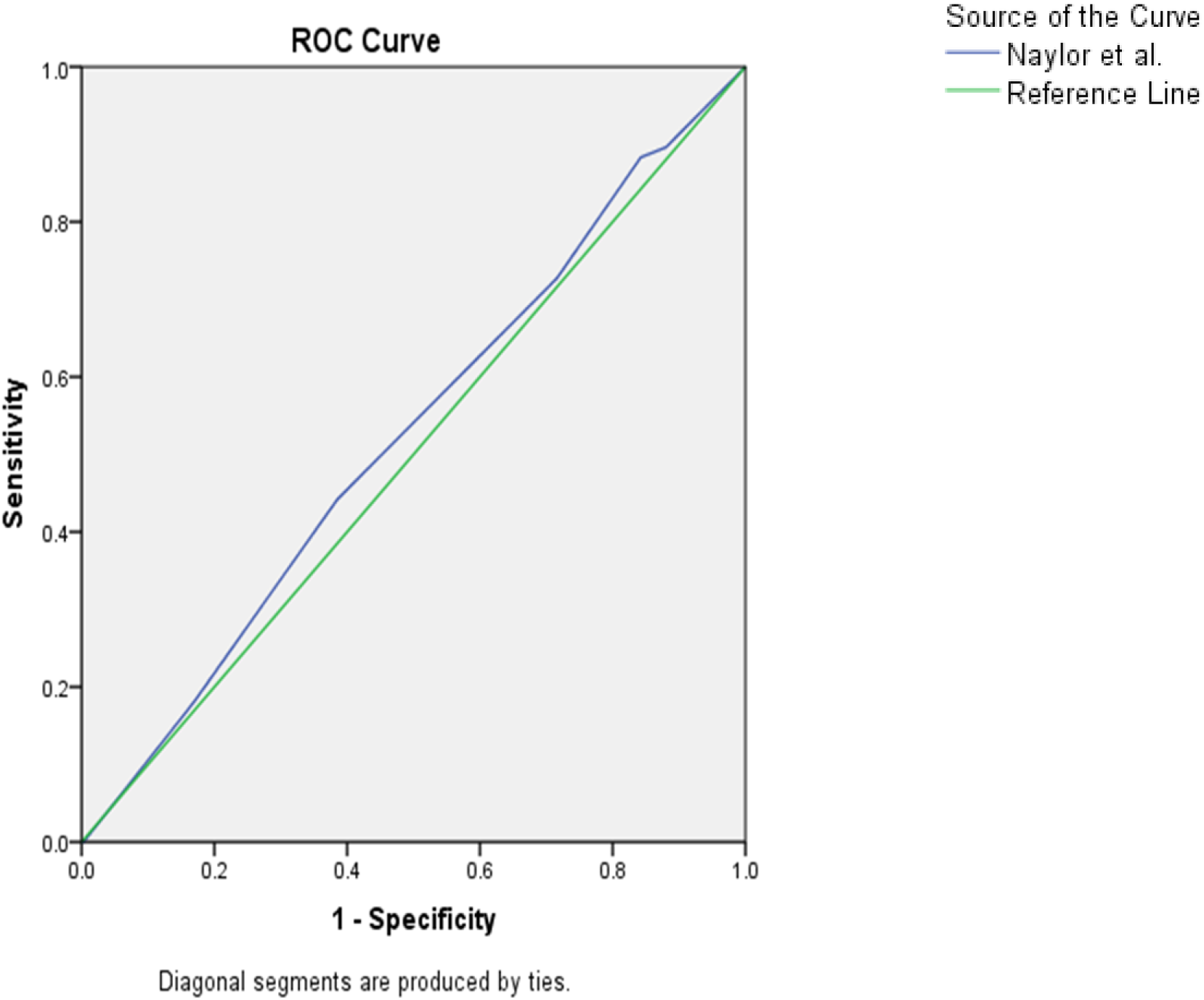
Receiver operating characteristic curves showing performance of Naylor *et al* clinical risk score in predicting GDM. Figure 1 shows the ROC curves for Naylor et al clinical risk score indicating performance of the test with reference to IADPSG diagnostic criteria. The Area Under the Curve (AUC) was 0.526 (95% CI 0.453-0.598) suggesting that the clinical risk score is non-discriminatory.

**Figure 2:**
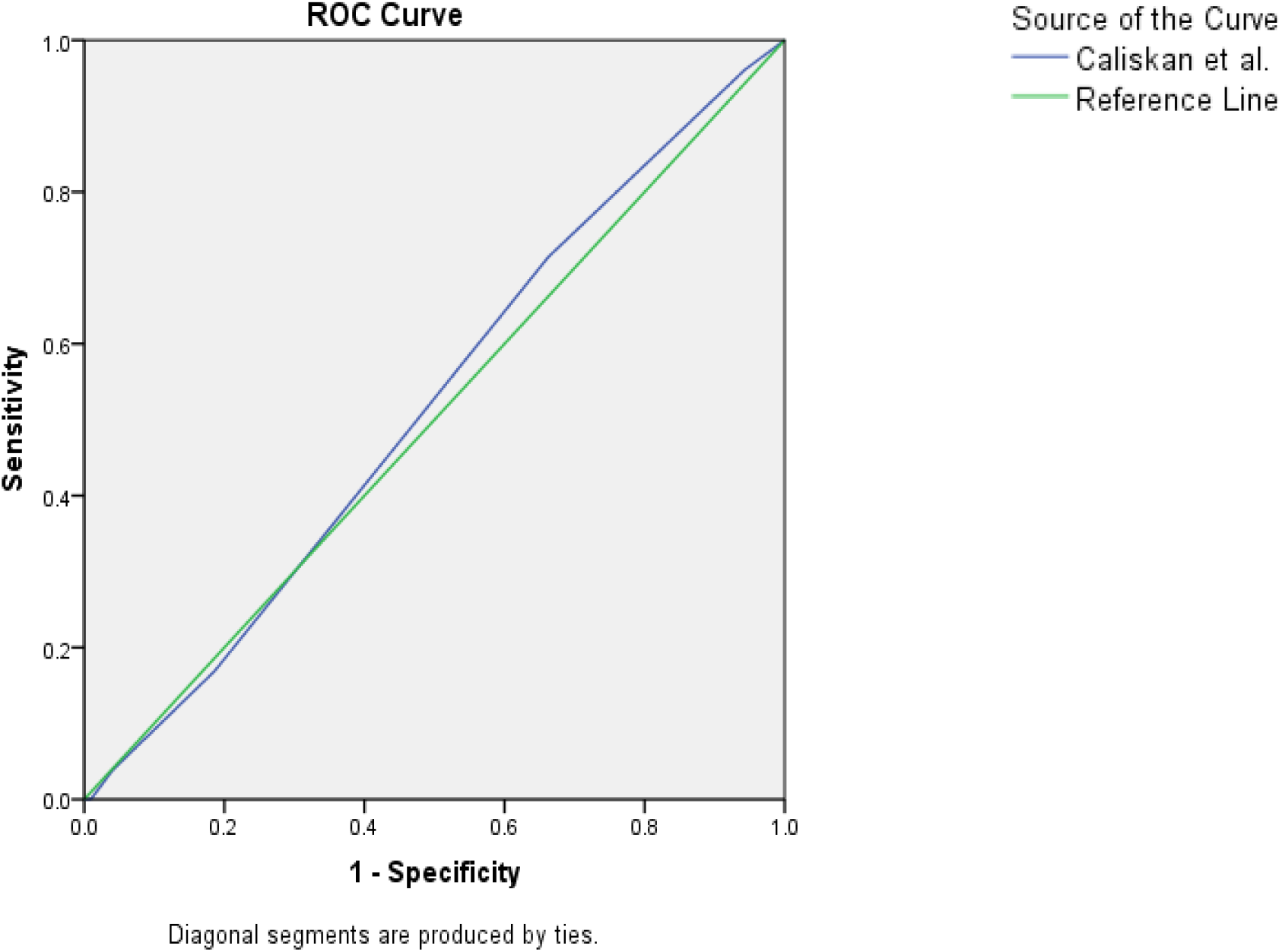
Receiver operating characteristic curves showing performance of Caliskan *et al* clinical risk score in predicting GDM. Figure 2 shows the ROC curves for Caliskan et al clinical risk score indicating performance of the test with reference to IADPSG diagnostic criteria. The Area Under the Curve (AUC) was 0.516 (95% CI 0.446-0.586) suggesting that the clinical risk score is non-discriminatory.

**Figure 3:**
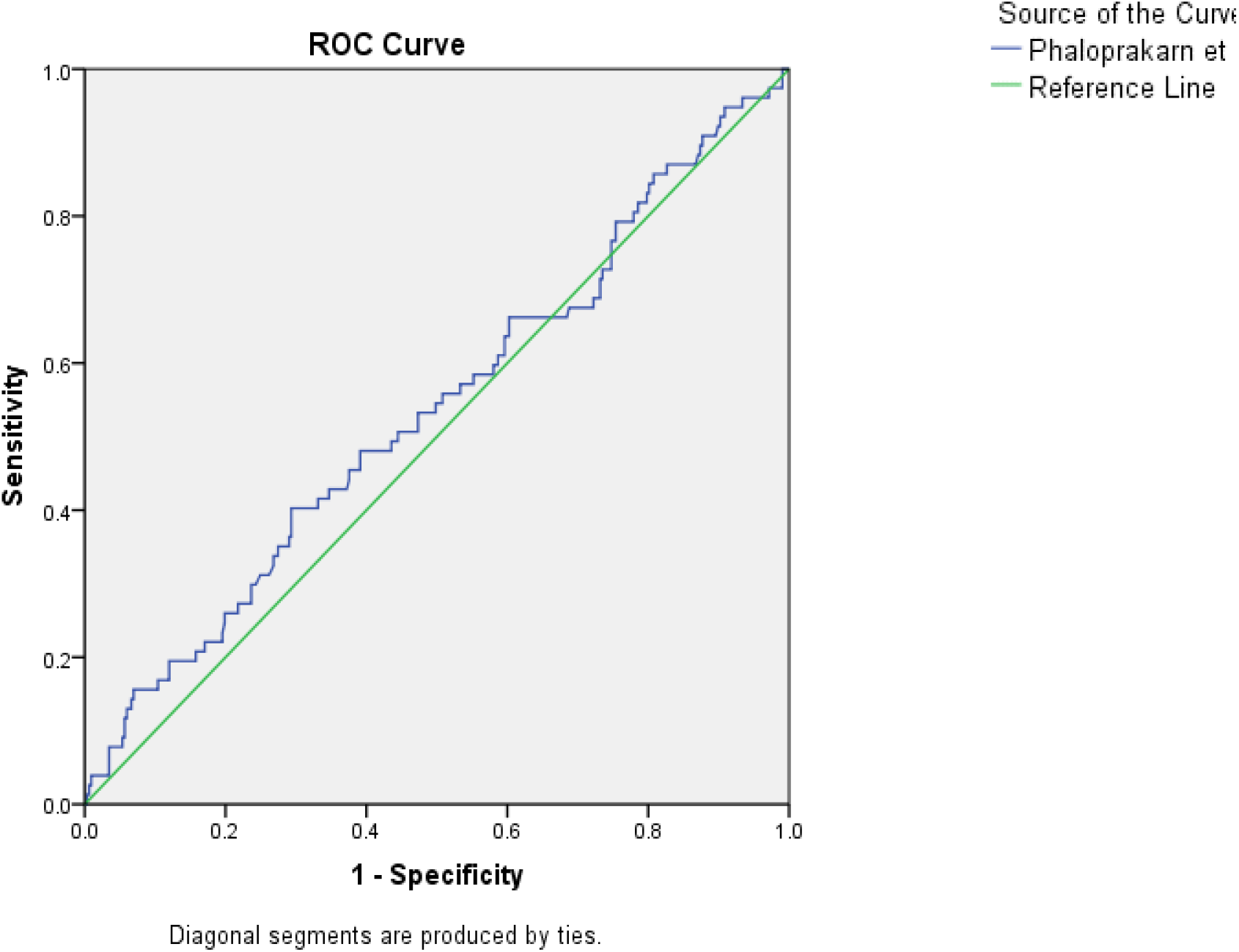
Receiver operating characteristic curves showing performance of Phaloprakarn *et al* clinical risk score in predicting GDM. Figure 3 shows the ROC curves for Phaloprakarn et al clinical risk score indicating performance of the test with reference to IADPSG diagnostic criteria. The Area Under the Curve (AUC) was 0.529 (95% CI 0.455-0.603) suggesting that the clinical risk score is non-discriminatory.

## Discussion

The prevalence of GDM among women in Lagos using the IADPSG/WHO 2013 criteria was 19%. This value is higher than the range of 1.5% to 11.5% obtained in earlier studies conducted in Lagos between 1987 and 2004.^20–22^ These studies used different diagnostic criteria and this may be responsible for the lower prevalence. The prevalence of GDM in this study is also higher than 4.9% reported in Ibadan by Fawole *et al* despite the fact that the study in Ibadan recruited only women that have high risk for GDM.^15^ It is possible that the higher prevalence of GDM in the index study may be due to the global increase in the prevalence of overweight and obesity over the years. However, the prevalence of GDM in this study was similar to 24% obtained in a recent study using the IADPSG criteria among pregnant women at the Lagos University Teaching Hospital notwithstanding that their method of glucose assay was not stated.^23^ The study by Onyenekwe *et al* in the South Eastern part of Nigeria used IADPSG criteria, but possibly overestimated the GDM prevalence at 35.9% due to the use of glucometer which could adversely affect the quality of plasma glucose measures and carrying out of OGTT only in those with risk factors for GDM.^7^ Although, the body of literature on the prevalence of GDM in Nigeria is large, the results varied due to the different diagnostic criteria and methods of glucose assay used by the researchers.

Comparing with studies conducted outside Nigeria that used the IADPSG diagnostic criteria, the prevalence of GDM was higher than the figures reported from United States of America (7.6%)^24^, Ireland (7.2%)^25^, and Turkey (14.5%)^26^, but lower than the prevalence of 34.9% and 38.6% for Punjab, India and Kuala Lumpur, Malaysia respectively^27, 28^. This is in keeping with previous findings that countries in North America and Europe have lower prevalence compared to sub-Saharan African countries while countries in the Middle East, North Africa and South East Asia have highest prevalence of GDM in the world. However, the prevalence of GDM in this study was similar to 18.3% reported from Marrakech and Safi districts in Morocco^29^. This could be a pointer to the rising burden of GDM in Nigeria. It is therefore important to ensure that all pregnant women are screened and diagnoses made for prompt treatment of the affected persons.

There are suggestions that the combination of clinical risk factors to form a clinical prediction model is associated with higher accuracy in detecting women with GDM rather than using each factor as an independent screening test. In this study, all the three established clinical risk factor models showed high sensitivity, low specificity, and poor accuracy for the detection of GDM. This means a substantial proportion of GDM patients will be detected by the models, but at the same time will not be a good diagnostic test for identifying those negative for GDM. Instructively, the three risk factor-based scoring models employed the two-step GDM screening strategy, which differs from the IADPSG/WHO guidelines and the practice in the index study. The stated aim for each of the models is not to replace the challenge-based glucose tolerance testing but to stratify low-risk women out of the second stage of screening. If our study participants were selectively screened based on Naylor, Caliskan, and Phaloprakarn risk scores, 15%, 32.8%, and 6.3% respectively would not have performed the diagnostic test. Out of this total, 85%, 83.2%, and 88% respectively would be negative if universal screening with diagnostic test was used. These high negative predictive values underline the fact that the risk factor-based models may be useful tools for screening patients out of the single-step 2-hour OGTT in our population.

The poor accuracy of the models is similar to the study by Adam and Reeder that tested eight clinical prediction models in the South African population including the three tested in this study and found that all the eighth models performed poorly in detecting GDM.^30^ The area under ROC for the three tested models in this study ranges from 51.6% to 52.9% and this is similar to 51.8% to 59.4% obtained in South Africa.^30^ These diagnostic accuracy is lower than the 73.3-83.2% reported for the derivation populations by the models. Out of the three clinical risk scores tested in this study, the Caliskan risk score had the highest accuracy. The Caliskan risk score was able to identify or predict correctly about 41% of the cases with GDM compared with about 30% and 24% for Naylor et al and Phaloprakarn et al risk scores respectively. The reasons for the low discriminatory power of these tests may be because they were derived from a different population and the fact that a selective screening approach and a different diagnostic criterion other than the IADPSG criteria were used in making these scoring systems.

In Nigeria, the guidelines for the management of GDM were developed by a team of endocrinologists only.^31^ The recommendations include risk assessment at booking, a one-step (75-g OGTT), or a two-step method (50-g GCT with 100-g OGTT) using C and C criteria for diagnosis. Despite the availability of this guideline on GDM, the practice varies across obstetric units in Nigeria. It is suggested that the risk models may be considered by obstetricians to screen out patients going for the diagnostic test. However, aside from saving cost and time, we must bear in mind the false negative cases and the known benefits of treatment. Further research to determine the accuracy measures of either a random blood test or the 50g glucose challenge as second-line screening tests to detect the false negative cases identified by the risk models warrants further consideration.

This index study has some limitations. First, the study used weight and height measured in early pregnancy for the calculation of body mass index unlike the risk factor-based models assessed in this study which used pregravid BMI of participants for computing their risk score. This may have affected the sensitivity and sensitivity of these clinical risk-based scoring models. In low-and middle-income countries, it is difficult to get a record of pre-pregnancy weight as most women do not go for preconception consultations in health facilities. Nonetheless, the influence of pregnancy on weight status at the early stage is considered minimal and as such measurement of weight and height and their association to compute BMI remains a valuable tool for the assessment of nutritional status in pregnancy. Second, most of the study subjects were of the high socio-economic class and this may have an influence on the prevalence of GDM. Lastly, this study was carried out in a tertiary health facility that receives high-risk referrals and therefore the study findings may not be representative of the general population and may also not be generalizable.

In conclusion, the burden of GDM is high in Lagos, Nigeria. This is in similarity to other resource-poor nations where available health facilities are grossly inadequate to take care of the existing problem of infectious diseases and the rising epidemics of non-communicable diseases. The clinical risk factor-based predictive models tested in this study could be used to select women for additional testing for GDM. In addition, efforts should be geared towards developing a cost-effective screening tool that is highly sensitive and at the same time more specific for the detection of GDM in low-resource settings.

## Data Availability

All relevant data are within the manuscript and its Supporting Information files.

## Acknowledgment

We appreciate and thank all the women that volunteered to participate in this study.

